# Clinical Validation and Diagnostic Utility of Optical Genome Mapping in Prenatal Diagnostic Testing

**DOI:** 10.1101/2022.05.11.22274975

**Authors:** Nikhil Shri Sahajpal, Ashis K Mondal, Timothy Fee, Benjamin Hilton, Lawrence Layman, Alex R Hastie, Alka Chaubey, Barbara R. DuPont, Ravindra Kolhe

## Abstract

The standard-of-care (SOC) diagnostic prenatal testing includes a combination of cytogenetic methods such as karyotyping, fluorescence *in situ* hybridization (FISH), and chromosomal microarray (CMA) using either direct or cultured amniocytes or chorionic villi sampling (CVS). However, each technology has its limitations: karyotyping has a low resolution (>5Mb), FISH is targeted, and CMA does not detect balanced structural variants (SVs) or decipher complex rearrangements in the genome. These limitations necessitate the use of multiple tests, either simultaneously or sequentially to reach a genetic diagnosis. This long-standing prenatal testing workflow demonstrates the need for an alternative technology that can provide high-resolution results in a cost and time-effective manner. Optical genome mapping (OGM) is an emerging technology that has demonstrated its ability to detect all classes of SVs, including copy number variations (CNVs) and balanced abnormalities in a single assay, but has not been evaluated in the prenatal setting. This retrospective validation study analyzed 114 samples (including replicates), representing 94 unique and well-characterized samples that were received in our laboratory for traditional cytogenetic analysis with karyotyping, FISH, and/or CMA. Samples comprised 84 cultured amniocytes, and 10 phenotypically normal and cytogenetically negative controls. Six samples were run in triplicate to evaluate intra-run, inter-run, and inter-instrument reproducibility. Clinically relevant SVs and CNVs were reported using the Bionano Access software with standardized and built-in filtration criteria and phenotype-specific analysis. OGM was 100% concordant in identifying the 101 aberrations that included 29 interstitial/terminal deletions, 28 duplications, 26 aneuploidies, 6 absence of heterozygosity (AOH), 3 triploid genomes, 4 Isochromosomes, 1 translocation, and revealed the identity of 3 marker chromosomes, and 1 chromosome with additional material not determined by karyotyping. Additionally, OGM detected 64 additional clinically reportable SVs in 43 samples. OGM demonstrated high technical and analytical robustness and a limit of detection of 5% allele fraction for interstitial deletions and duplications, and 10% allele fraction for translocation and aneuploidy. This study demonstrates that OGM has the potential to identify unique genomic abnormalities such as CNVs, AOHs, and several classes of SVs including complex structural rearrangements. OGM has a standardized laboratory workflow and reporting solution that can be adopted in routine clinical laboratories and demonstrates the potential to replace the current SOC methods for prenatal diagnostic testing. We recommend its use as a first-tier genetic diagnostic test in a prenatal setting.

## Introduction

Clinically recognized pregnancy loss and fetal anomaly (including birth defects and genetic disorders) occur in ∼15-25% and 3-5% of pregnancies, respectively; and represent a major public health concern, requiring morphological examination, biochemical screening, genetic counseling, invasive genetic testing, and appropriate perinatal management [**1-3**]. Chromosomal abnormalities account for 25-60% of the perinatal deaths and 15% of major congenital anomalies (diagnosed before 1 year of age), which are the leading cause of infant and childhood deaths [**4-6**]. For this reason, medical associations, including the International Society of Ultrasound and Obstetrics and Gynecology (ISUOG), the American College of Obstetrics and Gynecology (ACOG), and the American College of Medical Genetics and Genomics (ACMG), recommend that prenatal genetic testing be offered to all pregnant women, irrespective of the gestation and maternal age [**7-9**].

Typically, non-invasive prenatal screening (NIPS) is offered to pregnant women as early as 10-12 weeks of gestation [**10**], and invasive diagnostic testing is recommended to confirm the findings of the screening test or in pregnancies that have an abnormal ultrasound showing morphological defects. Diagnostic invasive testing may include chorionic villus sampling (CVS), amniocentesis, or periumbilical blood sampling (PUBS) performed at either 10-14 weeks or >15 weeks of pregnancy, respectively [**11**]. Historically, prenatal diagnostic testing has been offered since the 1960s using G-banded karyotyping, which enables genome-wide detection of structural variations (SVs) that include aneuploidies, unbalanced and balanced events (inversions, insertions, and translocations), and copy number variations (CNVs), at a resolution of ∼5-10 Mbp at best [**12**]. In the 1990s, fluorescence *in situ* hybridization (FISH) enabled targeted analysis and was implemented for rapid detection of the liveborn aneuploidies involving chromosomes 13, 18, 21, X, and Y using uncultured amniotic fluid cells [**13**]. The diagnostic yield with these conventional cytogenetic methods remained 3% in cases with positive biochemical screening, 6% in advanced maternal age, and 17-49% in fetuses with structural anomalies [**14**]. In the 2000s, chromosomal microarray (CMA) demonstrated the ability to detect sub-microscopic CNVs (micro-deletions and duplications) with a resolution of approximately 50-100 kbp and was recommended by ACOG and ISUOG as the first-line test in high-risk pregnancies with an abnormal ultrasound [**15,16**]. Although CMAs cannot detect balanced SVs or the orientation/location of duplications/insertions, they demonstrated an incremental diagnostic yield of 2.1-8.5% in fetuses with structural anomalies [**17**]. Recently, with the rapid progress in next-generation sequencing, exome sequencing has been evaluated as a diagnostic test and has demonstrated an incremental diagnostic yield of 19.4-27.4% in fetuses with structural anomalies [**17**]. However, because exome sequencing is limited to the detection of small variants (single nucleotide variants and small indels), cannot detect several classes of SVs, and has a large variant of unknown significance (VUS) burden, it is recommended only in select cases following a negative cytogenetic result [**17**]. Overall, the current diagnostic workflow relies on multiple technologies that are performed simultaneously or in a tiered fashion, often leading to increased cost and time to reach a diagnosis when clinical decision-making is time-limited. This situation warrants the evaluation of new technologies that have the potential to replace multiple technologies and improve the diagnostic yield in this important area of reproductive health.

In this context, optical genome mapping (OGM) is a next-generation cytogenomic technology with a resolution higher than SOC methods, and can detect all classes of SVs in a single assay. The technique is based on imaging ultra-long DNA molecules (>150 kbp) labeled at a specific 6 bp sequence motif (CTTAAG) that occurs across the entire genome (on average, every 5 kbp). Subsequently, the images are converted to molecules to generate a *de novo* genome assembly, which can be compared to a reference genome to identify germline SVs. The accurate and precise location of the labels enables the detection of different classes of SVs including deletions, duplications, and balanced/unbalanced genomic rearrangements including insertions, inversions, and translocations. Also, an independent coverage-based algorithm detects large CNVs and aneuploidies (similar to CMAs). Recently, several investigators have shown that OGM is 100% concordant when compared to standard-of-care (SOC) technologies in several settings including postnatal [**18**], hematological neoplasms [**19**], and solid tumors [**20**] in either a single site or multi-site studies. We have previously discussed the potential utility of OGM in prenatal diagnostic testing [**21**], but the application of the technology for a large-scale evaluation in a clinical setting for prenatal application remained elusive. Thus, in this clinical validation study (performed in a CLIA laboratory), OGM was compared to SOC methods (karyotyping, FISH and CMA), which included analyzing technical and analytical performance, reproducibility, and limit of detection for different SV classes. We report the feasibility and relative ease of implementing OGM for prenatal diagnostic testing compared to SOC methods with the platform demonstrating high robust technical and analytical performance and recommend OGM as a first-tier test in prenatal settings.

## Materials and Methods

### Sample selection

This retrospective validation study included the analysis of 114 samples (including replicates), representing 94 unique and well-characterized samples that were received in our clinical laboratory for cytogenetic analysis with karyotyping, FISH, and/or CMA. These comprised 84 prenatal samples (cultured amniocytes), of which, 15 were cultured in a T-75 flask and 69 in a T-25 flask. Of these 84 cultured samples, 25 were stored at -80°C as dry pellets (from 1-14 days before DNA isolation), and 59 were processed directly from culture (**Supplementary file 1**). Additionally, 10 phenotypically “normal” and cytogenetically “negative” samples (peripheral blood) were also analyzed to evaluate true negative/false-positive rates and calculate performance metrics. The OGM data were analyzed in a blinded fashion by the analyst (NS) and compared for concordance with SOC methods by Board-certified laboratory director (RK). The study was performed under IRB A-BIOMEDICAL I (IRB REGISTRATION #00000150), Augusta University. HAC IRB # 611298. Based on the IRB approval, the need for consent was waived; all PHI was removed, and all data were anonymized before accessing the clinical validation study.

### Optical genome mapping

Ultra-high molecular weight (UHMW) DNA was isolated, labeled, and processed for analysis on the Bionano Genomics Saphyr® platform following the manufacturer’s protocols (Bionano Genomics, San Diego, USA). Briefly, 0.6-1.5 M frozen cells or 0.4-1.5 M fresh cultured cells (**Supplementary file 1**) were digested with Proteinase K (PK) and lysed using Lysis and Binding Buffer (LBB). DNA was precipitated on a nanobind magnetic disk using isopropanol and washed using wash buffers A and B. The UHMW DNA bound to nanobind disk was eluted and quantified using Qubit broad range (BR) dsDNA assay kits (ThermoFisher Scientific, San Francisco, USA).

DNA labeling was performed following the manufacturer’s protocols (Bionano Genomics, USA), where 750 ng of UHMW DNA was labeled with DL-green fluorophores at a specific 6-base sequence motif (CTTAAG) using Direct Labeling Enzyme 1 (DLE-1) reactions. Following the labeling reaction, the DLE enzyme was digested using PK and the DL-green was removed using adsorption membranes. The DNA backbone was stained blue using DNA stain and quantified using Qubit high sensitivity (HS) dsDNA assay kits. Optical imaging was performed on the Saphry instrument by loading the fluorescently labeled DNA molecules onto flow cells of Saphyr chips. Analytical QC targets were set to achieve >160X effective coverage of the genome, >70% mapping rate, 13-17 label density (labels per 100kbp), and >230 kbp N50 (of molecules >150 kbp).

### OGM variant calling and data analysis

Genome analysis was performed using the de novo assembly pipeline included in the Bionano Access (v.1.6 or v.1/7)/Bionano Solve (v.3.6 or v.3.7) software for all the samples. Briefly, single molecules were used to generate de novo assembly of the genome, with the direct alignment of the consensus maps to GRCh38, reference human genome assembly, to detect germline SVs (Insertion, duplications, deletions, inversions, and translocations) based on the differences in the alignment of labels between the sample and the reference assembly. Additionally, a coverage-based algorithm enabled the detection of large CNVs and aneuploidies.

For data analysis, the variants were filtered using the following criteria: 1) the manufacturer’s recommended confidence scores were applied: insertion: 0, deletion: 0, inversion: 0.01, duplication: -1, translocation: 0, and copy number: 0.99 (low stringency, filter set to 0). 2) The GRCh38 SV mask filter that hides any SVs in difficult to map regions was turned off for analysis. 3) To narrow the number of variants to be analyzed, we filtered out polymorphic variants, i.e. those that appeared in >1% of an internal OGM control database (n>300). 4) The variants were further filtered by using a bed file containing genes/loci implicated in diseases/disorders, with the overlap defined as one label overlap with a 3kbp buffer corresponding to average label distance ± standard deviation around each gene/loci for SVs and 50kbp for CNVs.

### Performance metric evaluation

The performance was evaluated by calculating positive percentage agreement (PPA), negative percentage agreement (NPA), positive predictive value (PPV), negative predictive value (NPV), and accuracy. The data was analyzed in a blinded fashion (by NS) and compared for concordance with SOC methods (by RK) for concordance calculation, where a variant was considered concordant even if the size or breakpoint was slightly different. Differences in size and breakpoint are anticipated, as the resolution of OGM is higher than SOC methods. However, balanced SVs with centromeric breakpoints, or with a VAF <10% were excluded from the comparison (beyond the detection capabilities of OGM in the current iteration and with 160x coverage).

Additionally, we analyzed each case for all clinically relevant SVs, and report findings as “concordant” to previous SOC results and “additional findings with OGM” that were not reported with SOC methods. Although detailed investigation and confirmation of these additional findings remain beyond the scope of this manuscript, we have provided all clinically reportable SVs, and relevant literature in **Supplementary file 2** to assist future discoveries and consider these findings as likely real events, consistent with previous reports [**18-20**].

### Analytical comparison between OGM and SOC results

Of the 84 amniocytes samples, 26 samples were tested with karyotype and FISH, 21 with karyotype and CMA, 8 with CMA and FISH, 8 with karyotype, FISH, and CMA, 6 with only karyotype, and 15 with only CMA in the clinical setup (**Figure 1a**). Overall, the 84 cases harbored a total of 101 aberrations that included 29 interstitial/terminal deletions, 28 duplications, 26 aneuploidies, 6 absence of heterozygosity (AOH), 3 triploid genomes, 4 isochromosomes, 1 translocation, 3 marker chromosomes, and 1 additional material not identified by karyotyping (**Figure 1b**).

**Figure 1.**
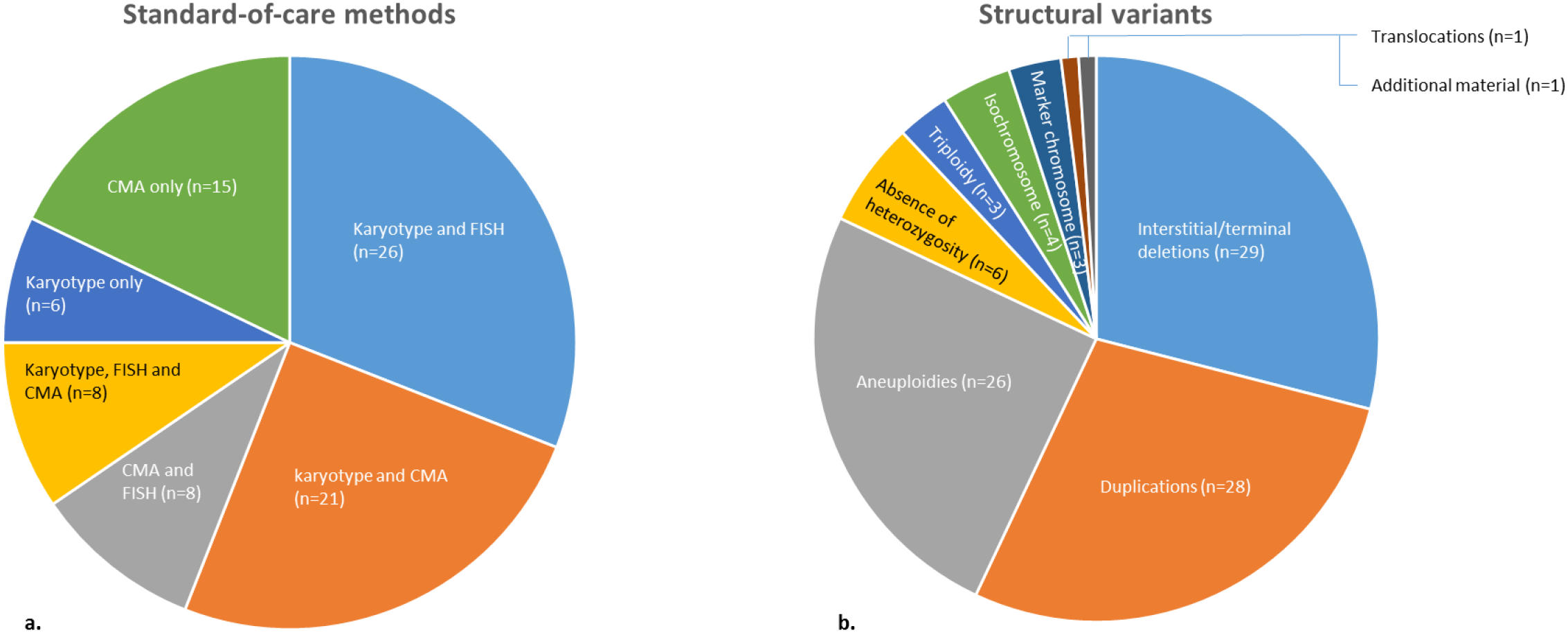
a) Standard-of-care methods used in the prenatal analysis for comparison in this study. b) Structural variants compared for concordance to optical genome mapping.

### Reproducibility studies

The reproducibility was evaluated by performing inter-run, intra-run, and inter-instrument comparisons. To evaluate inter-run and inter-instrument comparison, three samples were run in triplicate on three separate chips and two different instruments. To evaluate intra-run performance, three samples were run in triplicate on the same chip and instrument. The reproducibility was measured for both technical (QC metrics) and analytical (clinically reported variant) performance.

### Limit of detection studies

The OGM data can be processed using two pipelines, the de novo assembly pipeline, and the rare variant pipeline. The de novo assembly pipeline enables the detection of germline SVs and CNVs and is not designed for the detection of low mosaic events, while the rare variant pipeline compares individual molecules to a reference assembly enabling the detection of low mosaic events. To estimate the limit of detection (LoD) of the OGM platform, the samples in the LoD study were analyzed through both the de novo assembly pipeline and the rare variant pipeline with the same QC parameters and genome coverage of >160x. The LoD was assessed for various classes of SVs that included aneuploidy, translocation, interstitial deletion, and duplication. The LoD studies were performed for each variant class by diluting a sample with known variant and allele fraction (reported by SOC method) and diluting with wild type DNA to provide the following allele fractions 25%, 16.6%, 12.5%, 10%, and 5%. The samples at 12.5%, 10%, and 5% allele fractions (theoretical LoD range) were run in triplicate to confirm the LoD.

## Results

### Technical Performance: OGM quality control metrics and variant filtering

A typical run for OGM included the processing of six to fourteen samples in a batch for DNA isolation, and 12 samples for labeling, while three samples were loaded into one nanochannel array chip and two chips onto the Saphyr instrument at a given time. All 94 samples passed the quality control metrics and the 84 cultured amniocytes samples achieved an average N50 (>150 kb) of 305 kb (±31), map rate of 88.3% (±5.5), label density of 15.2/100 kb (±0.7), and average coverage of 226x (±27) (**Supplementary file 3**). The performance of samples stored at -80°C (1-14 days) was comparable to the samples processed immediately after culturing. Similarly, the performance of samples with a cell count of >1M was comparable to the samples with cell counts between 0.4 to 1 M (**Table 1**). In total, 496,586 SVs were identified in the 84 samples, with an average of ∼5,892 SVs per sample. Of all identified SVs, a total of 1,199 SVs remained after the filtration criteria were applied (99.98 % variants filtered out), with an average of ∼14 SVs per sample that were further interrogated for potential clinical relevance (**Supplementary file 4**).

**Table 1.**
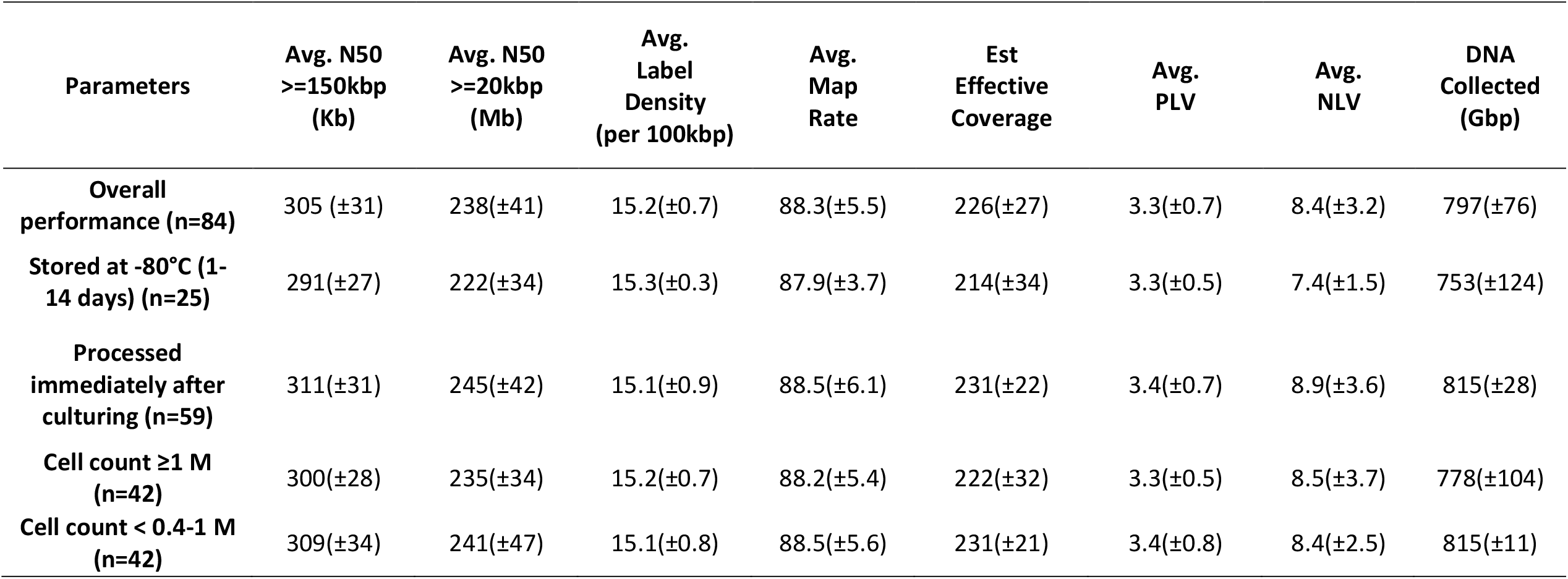
The performance of samples, overall performance, stored at -80°C (1-14 days), processed immediately after culturing, samples with a cell count of > 1M, and samples with cell counts between 0.4 to 1 M.

| Parameters | Avg. N50<br>≥150kbp<br>(Kb) | Avg. N50<br>≥20kbp<br>(Mb) | Avg.<br>Label<br>Density<br>(per 100kbp) | Avg.<br>Map<br>Rate | Est<br>Effective<br>Coverage | Avg.<br>PLV | Avg.<br>NLV | DNA<br>Collected<br>(Gbp) |
| --- | --- | --- | --- | --- | --- | --- | --- | --- |
| **Overall**<br>**performance (n=84)** | 305 (±31) | 238(±41) | 15.2(±0.7) | 88.3(±5.5) | 226(±27) | 3.3(±0.7) | 8.4(±3.2) | 797(±76) |
| **Stored at -80°C (1-**<br>**14 days) (n=25)** | 291(±27) | 222(±34) | 15.3(±0.3) | 87.9(±3.7) | 214(±34) | 3.3(±0.5) | 7.4(±1.5) | 753(±124) |
| **Processed**<br>**immediately after**<br>**culturing (n=59)** | 311(±31) | 245(±42) | 15.1(±0.9) | 88.5(±6.1) | 231(±22) | 3.4(±0.7) | 8.9(±3.6) | 815(±28) |
| **Cell count ≥1 M**<br>**(n=42)** | 300(±28) | 235(±34) | 15.2(±0.7) | 88.2(±5.4) | 222(±32) | 3.3(±0.5) | 8.5(±3.7) | 778(±104) |
| **Cell count < 0.4-1 M**<br>**(n=42)** | 309(±34) | 241(±47) | 15.1(±0.8) | 88.5(±5.6) | 231(±21) | 3.4(±0.8) | 8.4(±2.5) | 815(±11) |

### Analytical performance: 100% concordance with SOC results

OGM was 100% concordant in identifying the 101 aberrations reported with SOC methods (**Figure 1b**). Notably, 3 triploidy, 4 isochromosomes, 4 AOH regions, and 1 chromosome Y aneuploidy were manually inferred/called by the analyst (all variant classes are shown in **Figure 1b**). The ability to detect triploidy and AOH regions has only recently been added to the list of variants detected with OGM, in the Bionano Access software version 1.7. Thus, seven samples (three with triploidy and four with AOH regions) were analyzed using software v1.7. The detection capabilities and limitations of OGM (software v1.6 and v1.7) compared to SOC methods are provided in **Supplementary file 5**.

### Aneuploidy and copy number changes

Samples in this study included 26 aneuploidies, of which 9 were trisomy 13 (Patau syndrome), 6 were trisomy 18 (Edwards’ syndrome, **Figure 2a**), 4 were trisomy 21 (Down syndrome), and 7 were sex chromosome aneuploidies (**Figure 2b**). OGM was 100% concordant with SOC methods in detecting all aneuploidies, with all autosome and X chromosome aneuploidy called by the software, while, the chromosome Y aneuploidy in one sample had to be manually determined using the CNV visualization tool in the software (**Supplementary file 2**).

**Figure 2.**
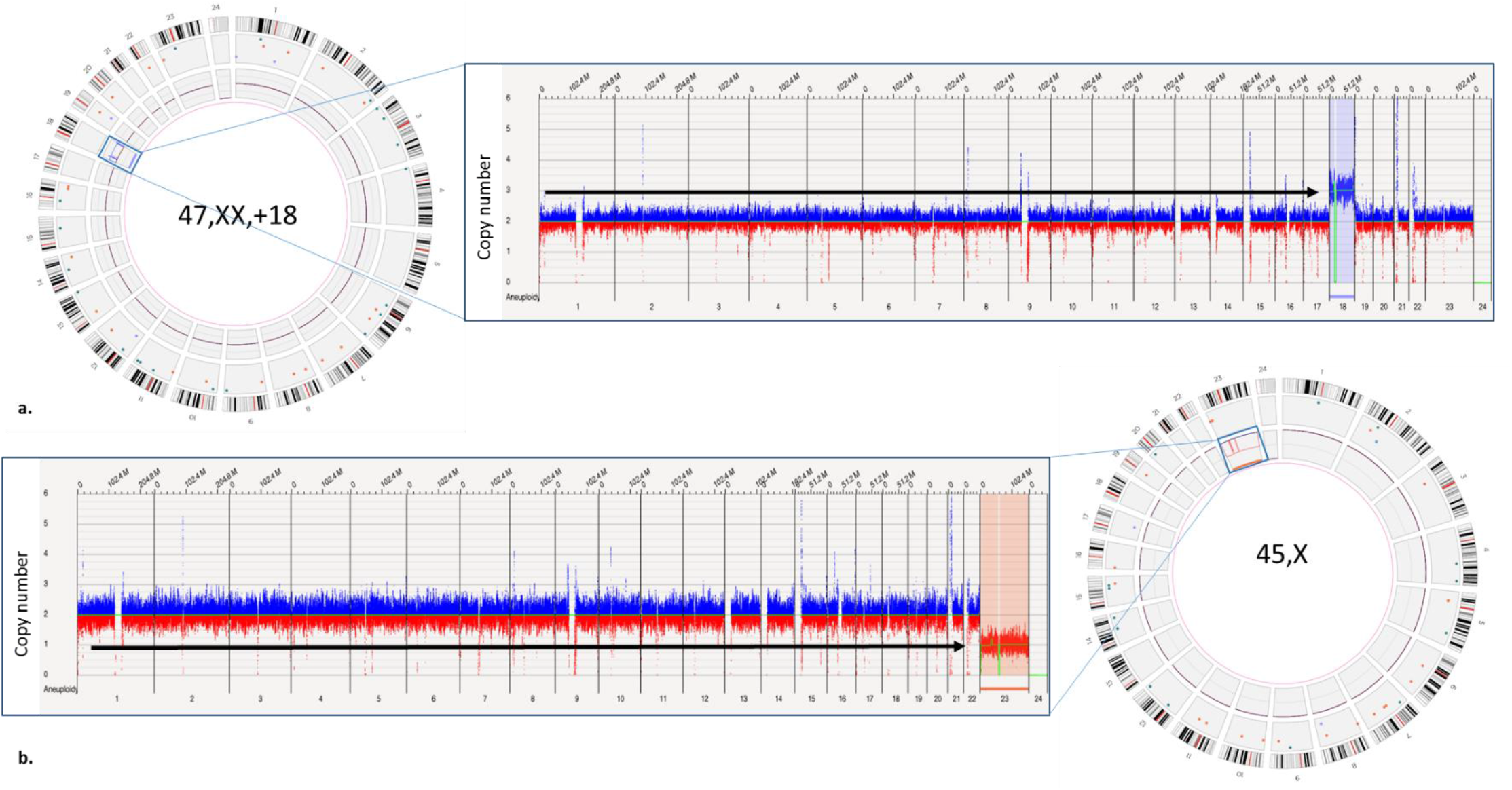
Representative example of aneuploidy detection with optical genome mapping. a) Left panel represents the circos plot with a copy number gain visible and highlighted with a blue box on the circos plot around the inner circle of the CNV track of chromosome 18, the right panel shows the whole genome CNV profile where the y-axis shows the copy number state and aneuploidy caller, and the x-axis shows the chromosome number. The copy number gain and aneuploidy call are observed on chromosome 18. b) The right panel shows the circos plot with a copy number loss visible and highlighted with a blue box on the circos plot around the inner circle of the CNV track of chromosome X, the left panel shows the loss of chromosome X to a CN state of 1 and an aneuploidy call observed for chromosome X on the whole genome CNV profile visualization.

The 29 deletions comprised of 27 interstitial and 2 terminal deletions that ranged from 95 kbp up to 16.7 Mbp and were automatically called by the software. The interstitial deletions larger than 500kb were called by both the SV and CNV algorithm, while deletions smaller than 500 kbp were only called by the SV algorithm as the threshold for the CNV algorithm was set at ≥500 kb. In addition, the terminal deletions were only called by the CNV algorithm (based on coverage), which is expected as the SV algorithm requires labels on both sides of the breakpoints of a deletion/duplication that could align to the reference assembly. The 28 interstitial duplications ranged from 132 kbp up to 18.2 Mbp and were called by the software. Similar to deletions, duplications below 500 kbp were called by only the SV algorithm (**Figure 3 a-d**).

**Figure 3.**
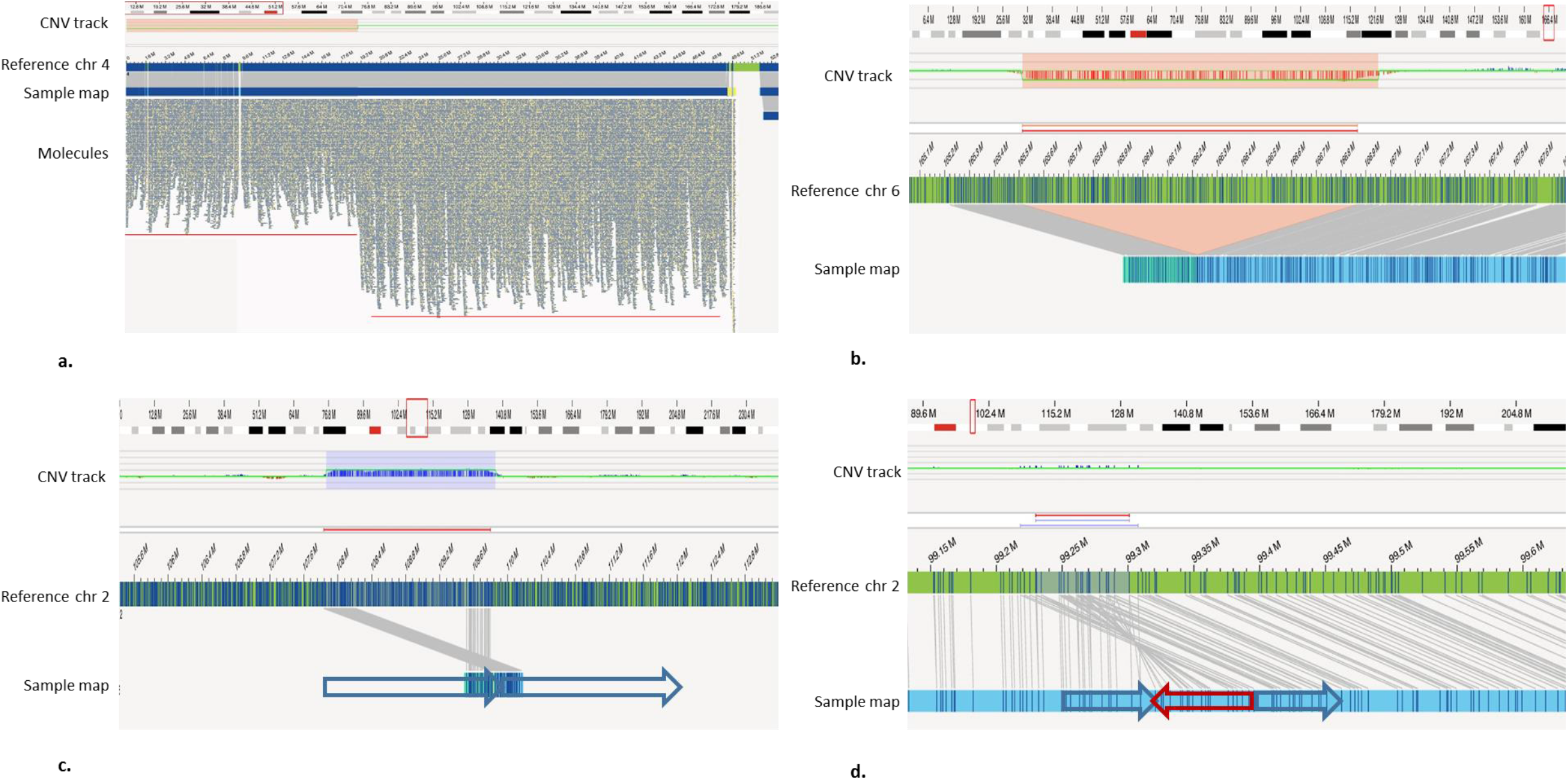
Representative examples of deletions and duplications detected with optical genome mapping. a) A terminal deletion 4p16.3p15.31(12985000_18938569)x1 detected with optical genome mapping, highlighted by the red block in the CNV track, which is supported by the lower coverage of molecules in the 4p16.3p15.31(12985000_18938569) region compared to remaining chromosome. b) Interstitial deletion highlighted in the CNV track and the SV track. c) A tandem interstitial duplication highlighted in the CNV and the SV track, where the orientation of the maps, represented with the blue arrows shows the orientation of the duplicated region of the genome. d) A small complex duplication, the orientation of the duplicated region is represented with the blue and red arrows.

### Isodicentric chromosome

The study included 4 samples with isodicentric chromosomes that included three isodicentric Y chromosomes and one isodicentric X chromosome. The isodicentric Y chromosomes were called upon manual interrogation, while the isodicentric X chromosome showed a complex CNV profile with inverted maps (inverted duplications called by the software), from which the isodicentric chromosome X was inferred (**Figure 4**). Notably, the isodicentric Y chromosome exhibits a characteristic mapping profile, which has been recently reported by Mantere et al, and can be visualized in the SV view of the software [**20**]. Consistent with the previous report, the three isodicentric Y chromosomes showed a genome map pattern where the maps were completely absent from the q arm starting from Yq11.222 compared to other XY samples. Notably, the Yq12 region has no coverage in all the samples, including controls as this region represents the repeat-rich gap in the reference genome (GRCh38, N-base gap), except for a 300 kb region at Yqter. However, the normal chromosome Y showed coverage up to Yq11.23, while the isodicentric Y chromosome showed coverage up to only Yq11.221. Two of the three sample showed inverted duplication maps (called by the software) at Yq11.221, which represent the fold-back fusion at the breakpoint and explain the formation of the isodicentric chromosome. Interestingly, in the recent report by Mantere et al, the software version 3.4/3.5 were unable to detect and call the inverted duplications at the breakpoint for the isodicentric Y chromosome [**20**], while we report that although the coverage pattern is the same, the software version 1.6 called the inverted duplications at the breakpoint in two of the three samples. We believe the breakpoint in the third sample where the inverted duplication was not detected is in the poorly mapped region where the reference assembly is incomplete.

**Figure 4.**
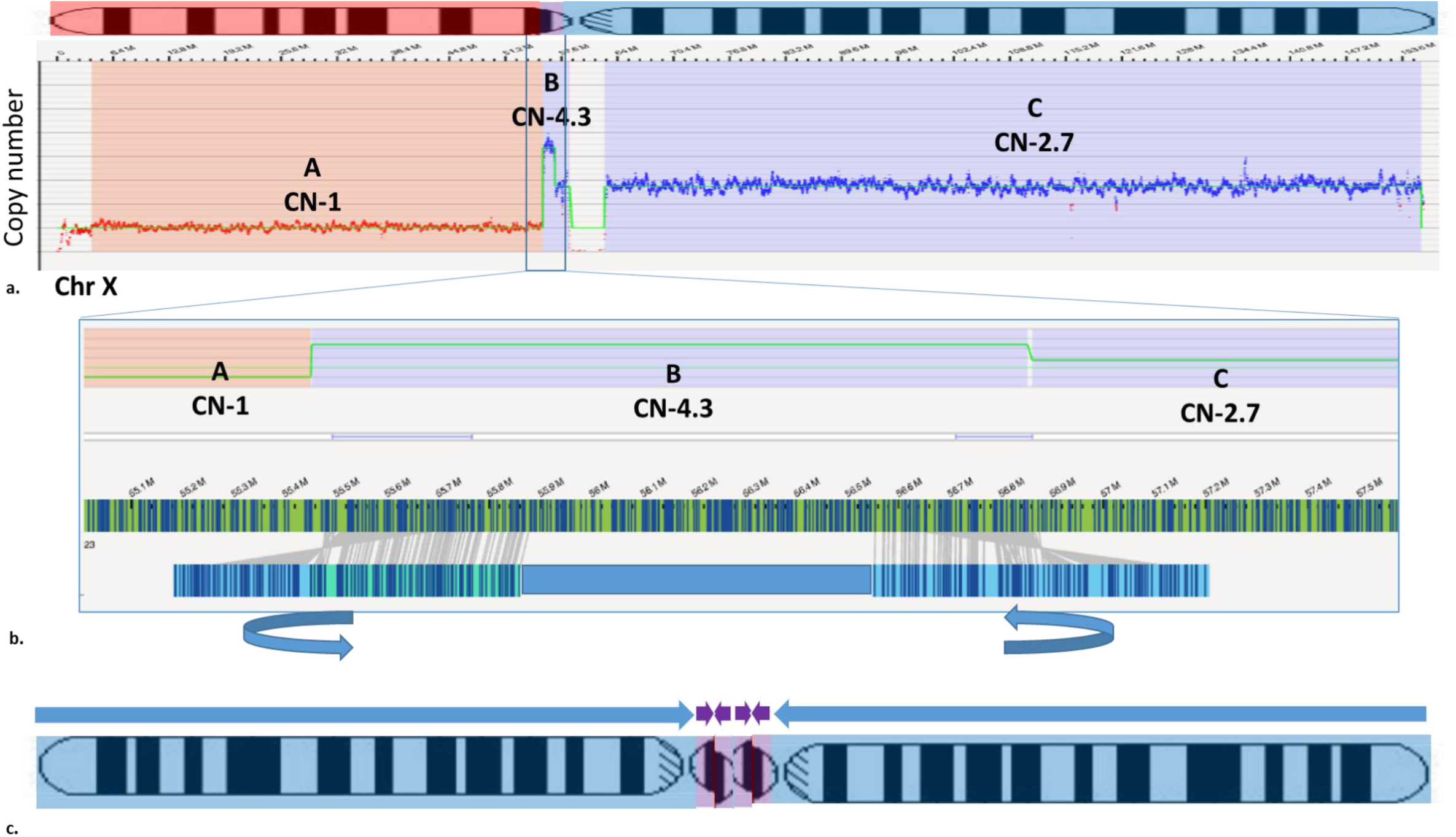
Derivation of isodicentric chromosome X from the optical genome mapping maps. a) The CNV profile of chromosome X is shown with three distinct regions with a CN state of 1, 4.3, and 2.7. b) The zoomed region highlighted in the blue box is represented with fold-back maps at the breakpoints of the region with a CN state of 4.3. c) The ideogram of the isodicentric chromosome X based on the CN state and map detected with optical genome mapping.

The isodicentric X chromosome demonstrated a complex CNV profile from bands Xp22.33 to Xp11.21 at a CN state of 1, a 1.3 Mbp region at Xp11.21 at a CN state of 4.3 with both breakpoints showing inverted maps (inverted duplications called by the software), and from Xp11.21 to Xqter at a CN state of 2.7. The ideogram showing isodicentric X chromosome was inferred from the CNV profile and inverted maps called by the software **(Figure 4**). Notably, the fractional CN state of 4.3 and 2.7 represent a mosaic event, which is consistent with the karyotyping results as the isodicentric chromosome was detected in 13 out of 30 cells.

### Triploidy and absence of heterozygosity (AOH)

This study included three samples with triploidy and four samples with AOH regions. These samples were analyzed with Bionano Access software version 1.7 since the ability to detect triploidy and AOH regions was not available in the previous version of the software. Of the three triploidy samples, two were 69, XXX and one was 69, XXY. The detection of triploidy was manually inferred from the variant allele fraction (VAF) track visualized in the circos plot and the whole genome view of the software. In a normal diploid genome, a homozygous SV is expected to have a VAF of 1 while a heterozygous variant is expected to have a VAF of 0.5, with a consistent normalized signal at 0.5, representative of the diploid genome (**Figure 5a**). In the triploid genome, the VAF signal is found to split into three distinct tracks at approximately 1, 0.66, and 0.33, as opposed to two tracks (**Figures 5 b and c**). The XXY is also evident from the copy number state of X and Y in this sample, the CN state of X is ∼1.5 and Y is ∼0.5 (**Figures 5 b and c**).

**Figure 5.**
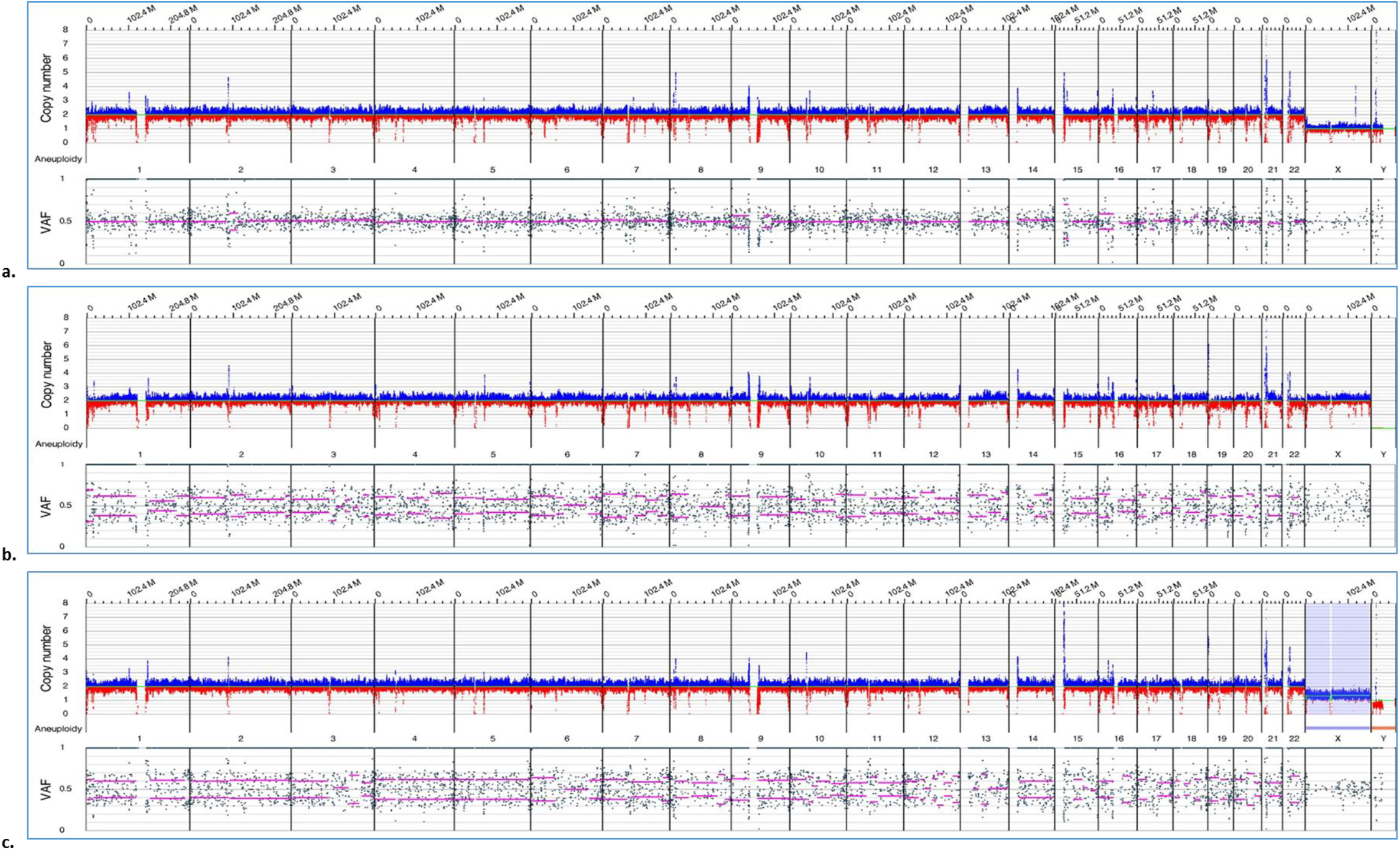
VAF patterns to identify the ploidy of the genome. a) In a normal diploid genome, a homozygous SV is assigned a VAF of 1 while a heterozygous variant is assigned a VAF of 0.5, with a consistent normalized signal at 0.5, representative of the diploid genome. b) In the triploid genome, the VAF signal is found to split into three distinct tracks as opposed to two tracks, and no copy number changes are observed in the CNV track for 69, XXX genome. c) In the 69, XXY genome, the VAF pattern is the same showing two distinct tracks, while the CN state of X is ∼1.5 and Y is ∼0.5.

In addition, three samples with five AOH regions ranging from 165 kbp to 64 Mbp, and one sample with ∼6.5% homozygosity of the autosomal genome were included in the study. The software called 2 of the 5 AOH calls automatically as the current software version can only detect AOH regions ≥25 Mbp. The software uses two algorithms i.e. the AOH algorithm and the VAF algorithm to detect the AOH regions. In the AOH algorithm, visualized as the AOH track in the whole genome visualization of the software, the SVs were plotted either as homozygous or heterozygous, with each SV represented with a dot at its coordinate throughout the genome. The regions of homozygosity were identified by a consistent decrease in heterozygous SV calls across a genomic region in the case sample compared to the level observed genome-wide in controls (**Figure 6**). After the blinded analysis, the 3 AOH regions <25 Mbp were manually evaluated and detected by the analyst using the same mechanism. The sample with ∼6.5% homozygosity was analyzed after removing the 25 Mbp filter and the software uniquely called 10 AOH regions comprising of ∼4.5% homozygosity of autosomal genome consistent with consanguinity. The removal of the size filter did not result in false positive AOH calls, as the calls were concordant with those detected with CMA. Notably, an average of 19 Mb (0.6%) of the genome is called as AOH regions by Bionano access software v1.7 in a subset of samples (n=10) when the default 25 Mbp minimum size filter is removed. Together, this demonstrates that the AOH tool can be used to detect consanguinity in the 6.5% range.

**Figure 6.**
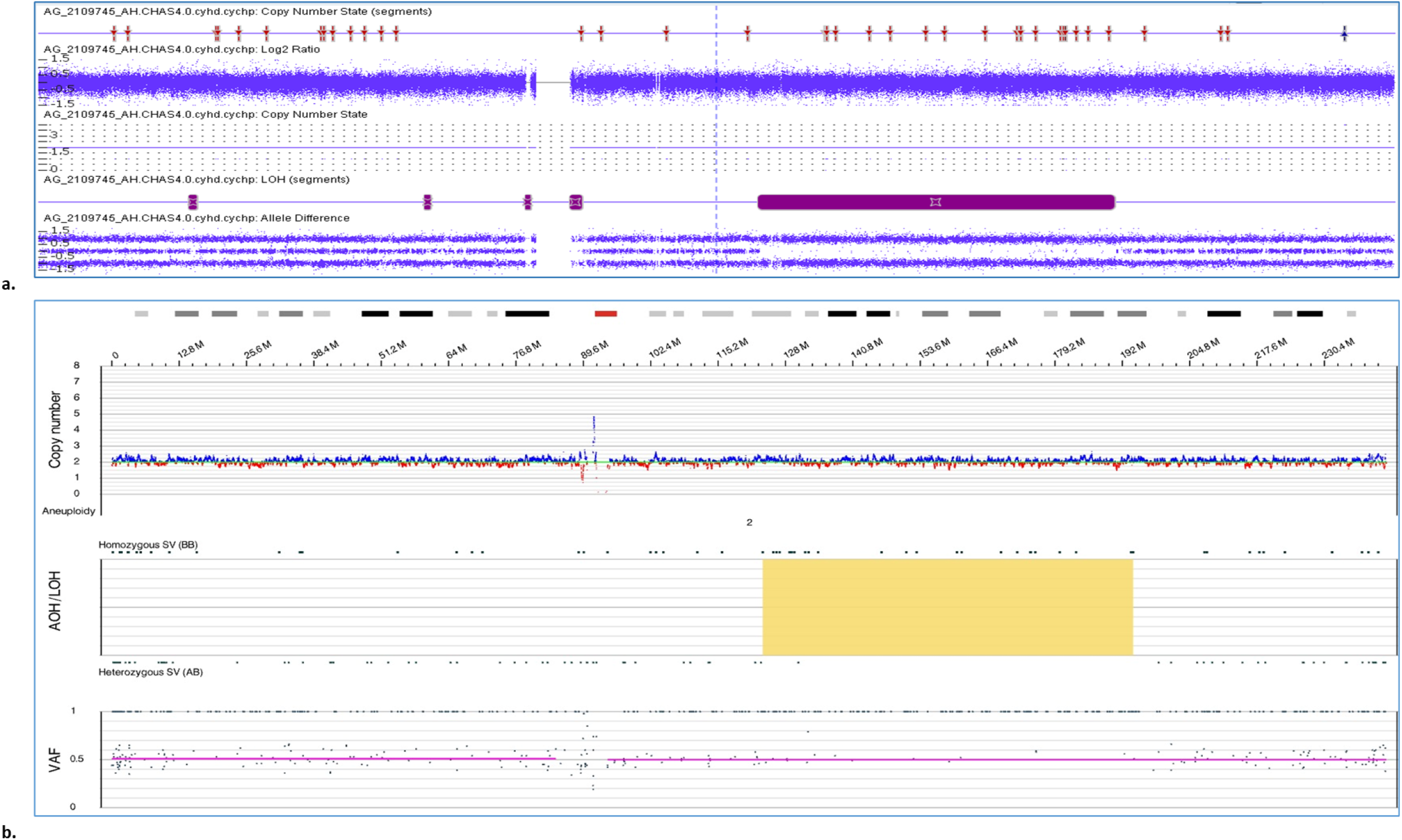
Detection absence of heterozygosity as observed with chromosomal microarray and optical genome mapping. a) Visualization of chromosome 2 showing the arr[GRCh37] 2q14.3q32.3(129125971_193128675)x2 hmz region. b) Visualization of the whole genome view showing the ogm[GRCh38] 2q14.3q32.3(123,674,249_194,059,050)x2 hmz AOH region highlighted in yellow.

### Additional findings and carrier screening

In addition to determining concordance, additional SVs were interrogated for relevance in these samples. Of the 84 samples, 43 samples harbored 64 SVs that were determined to be clinically reportable (including VUSs). Of these 64 SVs, 26 impacted genes with autosomal dominant (AD) inheritance, while 22 were in genes with autosomal recessive (AR) inheritance patterns. The disorder/syndromes included azoospermia, intellectual disability, Klippel-Feil syndrome 3, Spinocerebellar ataxia 1, and others (**complete list provided in Supplementary file 2**). Although confirmation of these additional findings is beyond the scope of this manuscript, we have listed all of the additional findings and relevant literature (OMIM IDs and inheritance patterns) for future investigation (**Supplementary file 2**).

### Performance metric evaluation

The performance metrics were calculated for 94 cases that included 84 prenatal cases and 10 negative cases of phenotypically normal controls. The 101 known aberrations detected with karyotyping/ FISH and CMA in the prenatal cases were used to calculate the true positive (TP) rate. OGM was concordant in detecting 101/101 aberrations. Of these 101 variants, 3 triploidy, 4 isochromosomes, 4 AOH regions, and one chromosome Y aneuploidy were manually determined/inferred and were not called by the software. As OGM has higher sensitivity/resolution and has been shown to detect additional variants compared to SOC methods, only the control samples were used for false positive (FP) and true negative (TN) calculations, where polymorphic variants were excluded and only pathogenic SVs (excluding autosomal recessive) were considered for calculation. No pathogenic SVs were detected with OGM in these negative cases, consistent with CMA results. From the 101 variants analyzed in this study, OGM was found to have a 100% sensitivity, specificity, positive predictive value (PPV), negative predictive value (NPV), and accuracy.

### Reproducibility studies

As part of this study, six samples were run in triplicate to evaluate inter-run, intra-run, and inter-instrument reproducibility. All 18 replicates passed the technical QC metric of >160x effective coverage of the genome, >70% mapping rate, 13-17 label density (labels per 100kbp), and >230 kbp N50 (of molecules >150 kbp) (**Supplementary file 6**). These six samples included three deletions, three duplications, and one aneuploidy. OGM detected these reported variants in each replicate, demonstrating a 100% inter-run, intra-run, and inter-instrument technical and analytical reproducibility (**Supplementary file 7**). In addition, the additional variants detected with OGM in these cases were also investigated and were found to be reproducible across the replicates, demonstrating high overall reproducibility.

### Limit of detection studies

The LoD studies were carried out to ascertain the lower LoD of the platform. The LoD was evaluated for the following variant classes: translocation, interstitial deletion, duplication, and aneuploidy with the two different analysis pipelines. With the denovo genome assembly pipeline, aneuploidy was detected down to 10% allele fraction, translocation to 12.5% allele fraction, and interstitial deletion and duplication to 16.6% allele fractions. With the rare variant pipeline, the interstitial deletion and duplication were detected consistently from 25% down to 5% allele fraction, whereas translocation and aneuploidy was detect down to 10% allele fraction. The aneuploidy was called by the software at 12.5%, but a lower aneuploidy threshold setting was used to call the aneuploidy at 10% allele fraction. Overall, these variants were detected in replicates at their respective LoD, with 5% allele fraction determined as the LoD for deletion and duplication, and 10% allele fraction for translocation and aneuploidy at 160x genome coverage with the rare variant pipeline (**Figure 7**).

**Figure 7.**
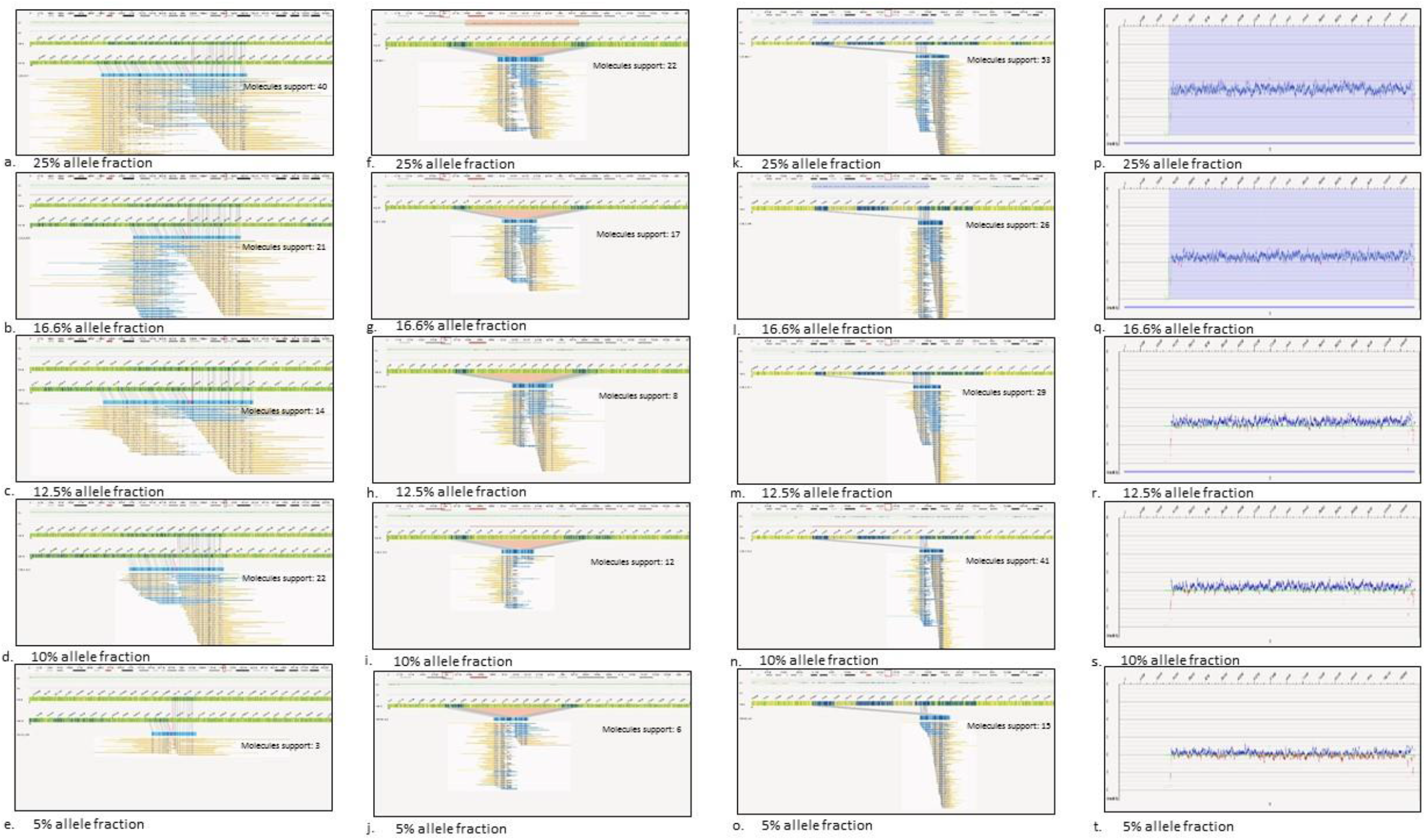
The limit of detection of optical genome mapping for aneuploidy, translocation, interstitial deletion, and duplication was assessed from 25% to 5% allele fraction and was found to be 5% allele fraction for interstitial deletion and duplication, and 10% for translocation and aneuploidy. a-e) Translocation: t(4;15)(122684318; 25010247) f-j) Interstitial deletion: 17p12p11.2(15752128_16669511)x1, k-o) Interstitial duplication: 2q12.3q13(108513707_110504318)x3, p-t) aneuploidy: trisomy 13.

## Discussion

Global medical associations such as ISUOG, ACOG, and ACMG recommend that prenatal genetic testing be offered to all pregnant women regardless of gestational and maternal age [**7-9**]. Invasive prenatal diagnostic testing is recommended after a positive NIPS test, in cases with abnormal ultrasound findings, or in high-risk pregnancies. As mentioned earlier, the SOC methods employed for prenatal diagnostic testing include, karyotyping, FISH, and CMA, which are performed either simultaneously or in a step-wise fashion to obtain a comprehensive cytogenetic analysis [**21**]. The requirement for multiple methods is evident from the SOC test results, as 75.2% of samples in our cohort were tested with at least two methods, while 9.4% required all three methods (karyotyping, FISH and CMA). This is primarily because each technology enables the detection of only certain types of SV classes (CMA) or is limited by targeted analysis (FISH) or resolution (karyotyping). To overcome the limitations of current SOC methods, there has been a significant interest in evaluating and validating OGM for prenatal diagnostic testing, as it detects all classes of SVs in a single assay. Herein, we discuss the results of our validation study performed as per CLIA requirements for a laboratory-developed test (LDT) and report the high technical and analytical performance of OGM with a 100% concordance to SOC methods.

### The technical performance of OGM

Sample processing in the prenatal workflow using cultured amniocytes is simple and standardized for the OGM assay. The first 15 samples in this study were cultured in T-75 flasks and the DNA was isolated using 1.5 M cells with 5 mm nanobind disks. However, with the availability of version 2 of the DNA isolation kits from the manufacturer that included 4 mm nanobind disks instead of 5 mm, we evaluated the feasibility of using T-25 flasks (using approximately 1 M cells) for DNA isolation. The samples cultured in T-25 flasks ranged from 0.4 M to 2.7 M cells, where several factors might have contributed to this wide range viz. the cells were pelleted for DNA isolation at different confluency (an attempt was made to harvest cells at >90% confluency), the different seeding volumes of the specimens, and the amniocentesis was performed at different gestational ages leading to differences in growth rates. Also, these were retrospective cases with cell aliquots stored (at different time points) in liquid nitrogen for several years, which might be another reason that could have accounted for these differences. However, all samples cultured in T-75 and T-25 flasks, ranging from as low as 0.4 M cells to 1.5 M cells passed the minimum QC metric and showed robust results with high molecule size, labeling density, and map rate. In addition, the cells stored at -80°C as dry pellets for 14 days did not affect the performance of the sample for OGM analysis (**Table 1**). These results demonstrate the feasibility of using OGM from a technical standpoint for prenatal diagnostic testing, as the culturing of cells in T-25 flasks is the standard protocol in cytogenetic laboratories for analysis and does not require any change to the culturing protocols in these laboratories. Furthermore, the storage of cell pellets at -80°C analyzed in this study allows the flexibility to store the cells and process them in batches for the OGM assay for maximum time and resource efficiency.

### Analytical performance of OGM compared to SOC methods (karyotyping, FISH and CMA)

OGM demonstrated excellent analytical performance, with 100% sensitivity, specificity, positive predictive value (PPV), negative predictive value (NPV), and accuracy in detecting 101/101 genetic aberrations in 84 prenatal samples. OGM has the unique ability to detect all classes of SVs including aneuploidies, deletions, duplications, balanced and unbalanced events (translocations, inversions, and insertions), AOH regions, and triploid genomes. This study included samples with multiple different SV types that are representative of the abnormalities observed in any clinical laboratory testing prenatal samples.

### Aneuploidies

Of the various SV types, aneuploidies are the most common aberrations detected in prenatal samples. Aneuploidies are the leading genetic cause of miscarriages, accounting for 29.4% when maternal age is <25 years, and 50% when maternal age is ≥40 years [**22**]. Furthermore, liveborn trisomy syndromes such as trisomy13 (Patau syndrome), 18 (Edwards syndrome), and 21 (Down syndrome) account for some of the most common birth defects. OGM was found to be 100% concordant in detecting the 26 aneuploidies, which included nine trisomy 13, six trisomy 18, four trisomy 21, and seven sex chromosome aneuploidies. Notably, one chromosome Y aneuploidy was manually called upon review, and we recommend a manual analysis of the CNV track, consistent with current routine practice in clinical laboratories performing CMA analyses.

### Deletions and duplications

OGM was 100% concordant in detecting the 29 interstitial/terminal deletions and 28 duplications. These deletions and duplications included microdeletions/duplications (<200kb) to large CNVs of 16.7 Mb and 18.2 Mb, respectively. The microdeletions/duplications that involve specific regions of the genome are associated with specific genetic diseases, which could only be detected with CMA in the current testing approach as the resolution of karyotyping is >5 Mb, and FISH panels do not offer genome-wide analysis. OGM detected all microdeletions/duplications and large CNVs using two different algorithms i.e. the SV and CNV algorithm. The use of two algorithms enables the detection of both small and large CNVs, as the SV algorithm is optimal for detecting small deletions/duplications up to 500/1500 kbp by matching and aligning labels in the sample to the reference assembly, while the CNV algorithm is optimal for detecting CNVs > 500 kb. The CNV algorithm utilizes coverage of labels to detect large CNVs and is uniquely capable of detecting CNVs in difficult to map regions such as the segmentally duplicated regions of the genome. In addition, the CNV algorithm enables the detection of terminal deletions/duplications, which are often associated with severe disorders. The two samples with terminal deletions, 1p36.33p36.32, and 4p16.3p15.31 included in this study were detected by OGM and are associated with chromosome 1p36 deletion syndrome (OMIM #607872) and Wolf-Hirschhorn syndrome [**23**], respectively.

### Isochromosomes

Isochromosomes have been reported in prenatal testing and are predominantly observed in advanced maternal age pregnancies [**24]**. The isodicentric Y chromosome has been associated with variable outcomes for which few reports have found it compatible with a normal male phenotype in absence of other abnormalities [**24,25]**, while other have reported cases with short stature [**26**] and mixed gonadal dysgenesis [**27]**. Similarly, isodicentric X chromosome has been reported with a normal female phenotype [**28]** and refractory anemia in a case of active isodicentric X [**29]**. Although the outcome remains variable, it is essential to detect these aberrations for accurate genotype/phenotype correlation and proper genetic counseling. The four isochromosomes included in this study were inferred from the mapping patterns visualized in the SV view of the software. The mapping patterns observed in our study are consistent with the recent results reported by Mantere et al. and represent a hallmark signature of the isodicentric Y chromosome with OGM [**18**]. The isodicentric X chromosome, or the isodicentric autosomes reported by Mantere et al., also show a characteristic CNV profile with fold-back maps that can be used by labs as a signature for isodicentric chromosomes [**20**]. We recommend the manual review CNV profile to consistently detect these SVs across laboratories using OGM.

### Triploidy and AOH

Triploidy affects 2 to 3% of pregnancies and accounts for approximately 20% of chromosomally abnormal first-trimester miscarriages. The three samples with triploidy were detected by the visualization of the unique signal in the VAF track in the “whole-genome view” of the software (**Figure 5**). The VAF and AOH tracks have only been included very recently in the software version 1.7, primarily to detect AOH regions. However, the VAF track demonstrates a distinct profile in triploid genomes that can be easily viewed in the “whole-genome view” of the software, and we recommend a manual review of all samples to detect or rule out triploidy in these samples.

Bionano Access version 1.7 software was used to analyze four samples with AOH regions ranging from 165 kbp to 64 Mbp. The recent updates in the software have utilized the existing SV zygosity status to detect AOH regions, and the results are encouraging for AOH regions >25 Mb. However, the software does not call AOH regions <25 Mb, but, we used manual review using the same mechanism and found the AOH regions with OGM concordant to CMA. Further work is required to develop and validate algorithms to call AOH regions <25 Mb. Since only 6 samples with AOH were included in this study, it was beyond the scope of this study to determine the sensitivity and specificity of AOH calling in regions <25 Mb. Given that on manual review these AOH regions were detected using the same mechanism used for calling AOH regions by the software, further investigation is required to determine the reproducibility and accuracy of AOH calling in regions less than 25 Mbp.

### Reproducibility and limit of detection

The OGM platform showed high reproducibility at the level of inter-run, intra-run, and inter-instrument, with all triplicates passing the QC parameters and all reported variants detected in respective samples, demonstrating the robustness of the assay. Reproducibility was also confirmed with the LoD studies, where the four variants were consistently detected in triplicate at 12.5%, 10% allele fractions, and deletion/duplication at 5% allele fraction. This is the first study to formally assess the LoD, which was found to be 5% allele fraction for interstitial deletions and duplications, and 10% allele fraction for translocation and aneuploidy, at ∼160x genome coverage with the rare variant pipeline.

### Limitations of the Study

The study had a few limitations, which include that only the cultured amniocytes were analyzed with OGM, while other sample types such as CVS and direct amniocytes/CVS were not analyzed. Notably, CVS samples have been previously evaluated with OGM, and have been demonstrated to perform optimally on the OGM technology [**18,21**]. Given that this study establishes the validity of OGM in detecting the different SV classes in the prenatal settings, a next logical step could be to determine if direct amniocytes could be used to perform OGM to reduce the TAT but this was beyond the scope of the current study.

### Recommendations

Since OGM can detect the different classes of SVs and alleviates the need for multiple tests (karyotyping and CMA), we recommend that after a negative STAT FISH test, that sample should be tested with OGM instead of karyotyping and/or CMA. The turn-around time (TAT) with OGM for analysis in the prenatal setting would be 10-14 days from sample collection to reporting (7-10 days to culture T-25 flask + 4 days for OGM), which is comparable to testing with karyotyping/CMA, or sequential testing with karyotyping and CMA. We are interested to see if OGM can be performed on direct amniocytes (beyond the scope of current work), which would alleviate the need for culturing (culturing would still be performed for back-up, a common practice in clinical laboratories), and can greatly reduce the TAT. In cases negative on OGM, karyotyping could be performed as a reflex test to rule out/ rule in balanced Robertsonian translocations.

In conclusion, this study demonstrates the robustness of the OGM assay for invasive prenatal diagnostic testing. This study demonstrates the unique ability of OGM to detect all classes of SVs including triploidy, aneuploidies, microdeletion/duplications, isochromosomes, AOH regions, balanced and unbalanced events, and complex structural rearrangements in a single assay. The higher resolution achieved with OGM can play a significant role in prenatal care and management as a next-generation cytogenomic tool. The standardized laboratory workflow and reporting solution with no custom bioinformatics requirements makes it suitable for adoption by clinical laboratories, and we recommend OGM as a potential first-tier test in prenatal settings.

## Data Availability

All relevant data is included in the manuscript and supplementary files

## Ethics approval

The study was approved by the IRB A-BIOMEDICAL I (IRB REGISTRATION #00000150), Augusta University. HAC IRB # 611298. Based on the IRB approval, the need for consent was waived; all PHI was removed, and all data was anonymized before accessing for the study.

## Declarations

RK has received honoraria, and/or travel funding, and/or research support from Illumina, Asuragen, QIAGEN, Perkin Elmer Inc, Bionano Genomics, Agena, Agendia, PGDx, Thermo Fisher Scientific, Cepheid, and BMS. NSS owns limited number of personal stocks of Bionano Genomics Inc. AH, and AC are salaried employee at Bionano Genomics Inc. All other authors have no competing interests to disclose.

